# Medical ChatGPT – A systematic Meta-Review

**DOI:** 10.1101/2024.04.02.24304716

**Authors:** Jan Egger, Malik Sallam, Gijs Luijten, Christina Gsaxner, Antonio Pepe, Jens Kleesiek, Behrus Puladi, Jianning Li

## Abstract

Since its release at the end of 2022, ChatGPT has seen a tremendous rise in attention, not only from the general public, but also from medical researchers and healthcare professionals. ChatGPT definitely changed the way we can communicate now with computers. We still remember the limitations of (voice) assistants, like Alexa or Siri, that were “overwhelmed” by a follow-up question after asking about the weather, not to mention even more complex questions, which they could not handle at all. ChatGPT and other Large Language Models (LLMs) turned that in the meantime upside down. They allow fluent and continuous conversations on a human-like level with very complex sentences and diffused in the meantime into all kinds of applications and areas. One area that was not spared from this development, is the medical domain. An indicator for this is the medical search engine PubMed, which comprises currently more than 36 million citations for biomedical literature from MEDLINE, life science journals, and online books. As of March 2024, the search term “ChatGPT” already returns over 2,700 results. In general, it takes some time, until reviews, and especially systematic reviews appear for a “new” topic or discovery. However, not for ChatGPT, and the additional search restriction to “systematic review” for article type under PubMed, returns still 31 contributions, as of March 19 2024. After filtering out non-systematic reviews from the returned results, 19 publications are included. In this meta-review, we want to take a closer look at these contributions on a higher level and explore the current evidence of ChatGPT in the medical domain, because systematic reviews belong to the highest form of knowledge in science.

## Introduction

Comprehensive systematic reviews that synthesize the findings of multiple studies can provide a more robust and reliable understanding of ChatGPT’s performance, its limitations, and the challenges that still need to be addressed, which are particularly useful in fields where research is abundant and rapidly evolving, such as the adoption of ChatGPT in healthcare. They allow researchers to quickly grasp the key aspects of a topic without having to sift through numerous individual studies. The increasing popularity of ChatGPT among healthcare professionals and researchers alike leads to a surge in research focusing on the evaluation of ChatGPT’s performance in different application scenarios, such as medical consultation [1], research [2], education [3], or different medical specialties, such as neurology [4, 5], pediatric [6, 7], cosmetic surgery [8, 9] and dermatology [10, 11]. Each of these fields presents unique challenges and opportunities for ChatGPT. Understanding how ChatGPT performs in these different contexts is crucial for its continued development and improvement.

Due to the large amount of publications produced in a relatively short time, it is demanding for researchers to stay up-to-date with the latest development of ChatGPT in their specific domain and understand its challenges and limitations. Systematic reviews provide a quick and informative overview on the use of ChatGPT in a particular scenario or specialty [12, 13, 14, 15, 16, 17, 18, 19], which keep researchers updated with the field.

A meta-review on ChatGPT in healthcare synthesizes the findings of multiple systematic reviews, and therefore takes a broader view of the field, going beyond the scope of systematic reviews that focus only on specific scenarios or specialities. It aims to provide a concise and high-level summary of the current state of ChatGPT in the general healthcare sector [20], and help researchers, healthcare professionals, and policymakers understand the big picture in order to make sound decisions and policies. Future research directions can also be suggested.

We position our work as a meta-review of systematic reviews of ChatGPT in healthcare, which look at systematic reviews across various application scenarios and medical specialties. The aim of the meta-review is to provide a concise yet comprehensive summary of the status quo of ChatGPT in the healthcare sector, assess the overall performance of ChatGPT, identify common trends and patterns, highlight key challenges and limitations, and point out areas where further research is needed. However, it is important to notice that, at this stage, the number of comprehensive systematic reviews is still small, and the majority of related publications remain to be high-level commentaries or small-scale evaluations [21], which highlights a gap in the literature and an opportunity for future research.

## Meta-Review of Systematic Reviews

### Methodology

As search strategy, we used the search string “*ChatGPT*” in PubMed and restricted the “*Article type*” to “*Systematic Review*”, and ordered the resulting publications by date in ascending order:

https://pubmed.ncbi.nlm.nih.gov/?term=“ChatGPT”&filter=pubt.systematicreview&sort=date&sort_order=asc

Afterwards, the resulting publications were screened by two authors (J.E. and J.L.). The inclusion criteria were systematic reviews about ChatGPT in healthcare, also if they have been systematic scoping reviews, like [29]. The exclusion criteria were non-systematic reviews, publications that don’t cover mainly a healthcare topic, comments, editorials and publications that were not written in English. The overall search strategy used in this systematic meta-review is shown in Diagram 2. and utilizes a slightly adapted version of the PRISMA Flow Diagram.

**Diagram 1.**
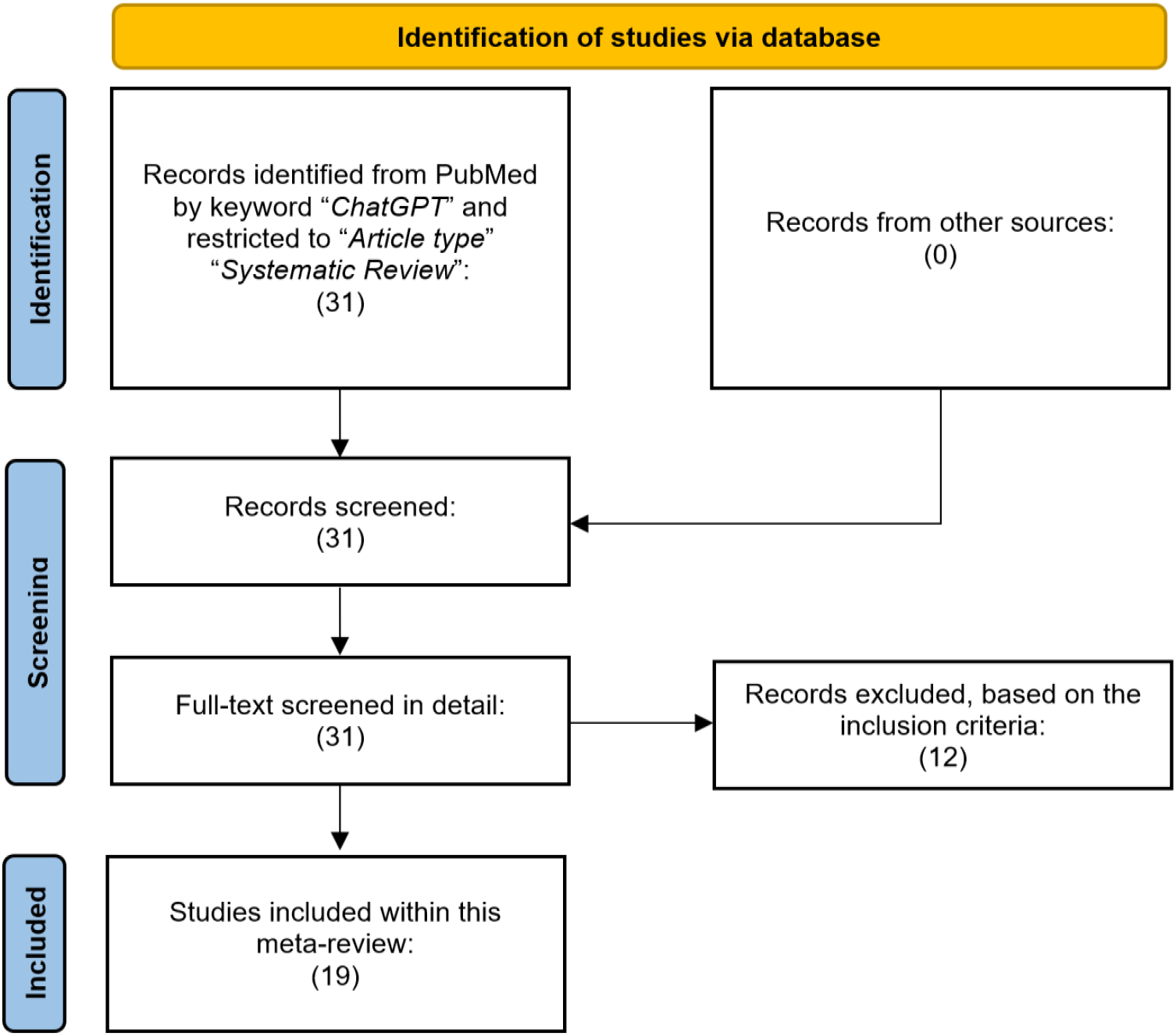
The overall search strategy used in this systematic meta-review utilizing a slightly adapted version of the PRISMA Flow Diagram.

Table 1. summarizes the published systematic reviews about ChatGPT in the medical domain according to PubMed (status as of March 2024), which are arranged by epub (electronic publication) date in chronological order. The earliest and most representative publication is from Sallam M. [22], which was published already online on the 19th of March 2023, so exactly one year ago from the systematic search for this contribution. It covers a variety of publications that apply ChatGPT to the healthcare domain, whereby several of them are more on an abstract level, like comments or editorials, e.g. [24]. There were recently some critics that the publication does not fulfill all criteria for a systematic review [25] with a response from Sallam [26]. However, the outcome of the review showed strong evidence that the prospect of ChatGPT in healthcare is rather promising.

**Table 1.** List of published systematic reviews about ChatGPT in the medical domain according to PubMed (status as of March 19 2024); ordered by epub (electronic publication) date.

| **Publications** | **Date (epub)** | **Num. Refs** | **Outcomes** |
| --- | --- | --- | --- |
| Sallam M. [22] | 19 March 2023 | 96 | The use of ChatGPT in healthcare education and research is inevitable. However, substantial research is needed to evaluate ChatGPT's performance and impact in practice. |
| Tustumi T. [23] | 8 May 2023 | 25 | Even though the use of ChatGPT in medicine is promising, its limitations must be carefully addressed. Responsible use of the tool must also be observed by healthcare professionals. |
| Anghelescu A. [12] | 2 June 2023 | 43 | ChatGPT is currently not fully capable of conducting critical systematic reviews. Inconsistency is observed by comparing the information extracted from a conventional system review and ChatGPT in terms of the treatment effects of Actovegin/AODEJIN on ischemic stroke. |
| Tiwari A. [13] | 13 June 2023 | 21 | In public health dentistry, ChatGPT cannot replace dentists, who not only make diagnoses but also analyze clinical findings, even though ChatGPT is a powerful tool for dentistry. |
| Balla Y. [14] | 15 July 2023 | 42 | The use of ChatGPT in pediatrics medicine has met with both challenges and opportunities. |
| Roman A. [15] | 15 August 2023 | 32 | ChatGPT has the potential to improve diagnosis, treatment and patient outcome in neurosurgeries, while a responsible and ethical use of the AI tool must be followed. |
| Levin G. [27] | 21 August 2023 | 6 | ChatGPT shows overall passing grade in medical examinations. |
| Garg R. K. [16] | 11 September 2023 | 145 | ChatGPT is helpful as an assistant in research and scientific writing, whereas issues concerning its accuracy, authorship and bias need to be further addressed. |
| Bečulić H. [18] | 14 October 2023 | 49 | More research is needed to understand the benefits and limitations of using AI tools such as ChatGPT in neurosurgery. Currently they should be used with caution. |
| Nikolas S. [36] | 28 November 2023 | 41 | ChatGPT depicts promising performance in various clinical tasks. Following studies should focus on expanding the application scope of ChatGPT and investigating the solutions for ethical issues. |
| Hiroj B. [35] | 29 November 2023 | 51 | ChatGPT can give appropriate answers to dentistry-related questions, which however need to be carefully evaluated before adoption. |
| Eleni M. [34] | 10 December 2023 | 158 | Smart digital tools like ChatGPT can be useful for mindfulness training and mitigate mental health issues. |
| Eyal K. [33] | 25 December 2023 | 13 | ChatGPT has shown potential in gastroenterology, especially in terms of helping with the interactions between patients and physicians. However, customization and ethical issues should be further addressed. |
| Hussain A. Y. [32] | 4 January 2024 | 93 | ChatGPT has shown transformative potential for a variety of medical applications. Responsible and ethical use of the tool must be observed. |
| Li J. [21] | 15 January 2024 | 147 | The development of ChatGPT in healthcare should aim for the respective medical speciality, where a unified evaluation system is still lacking. |
| Hugo C. T. [31] | 19 January 2024 | 37 | ChatGPT demonstrates potential in improving the efficiency and diagnosis accuracy of radiological workflows, as well as in assisting research. An interdisciplinary team involving AI experts, radiologists and policymakers are needed for the implementation of ChatGPT in radiological practice. |
| Wei Q. [30] | 8 March 2024 | 86 | The current scheme for evaluating ChatGPT's ability to answer medical inquiries are heterogeneous and lack conformity in reporting. Future studies are needed to objectively quantify ChatGPT's performance in medical question and answering. |
| Xiaojun X. [29] | 15 March 2024 | 55 | ChatGPT can potentially transform medical education. Special attention should be given to how ChatGPT affects the students and teachers. |
| Anusha S. [37] | 13 March 2024 | 30 | ChatGPT shows overall satisfactory performance in various medical examinations containing different questions types. Its performance drops as the difficulty of the question increases. |

For some publications, the application of ChatGPT in a specific medical speciality or application scenario is discussed, including ischemic stroke [12], dentistry [13], pediatrics [14], neurosurgery [15, 18], patient care [16], medical education and examination [17, 18, 27] and scientific writing [19, 24]. In particular, Najafali D. et al. [19] discussed whether ChatGPT is able to independently carry out a systematic review on cosmetic surgery. It is concluded that the content generated by ChatGPT is not rigorously following the Preferred Reporting Items for Systematic Reviews and Meta-Analyses (PRISMA) guidelines for systematic reviews, and that in its current form, ChatGPT can be beneficial for exploring systematic review ideas. Chen TJ. et al. [24] conclude that ChatGPT and other artificial intelligence (AI)-based language tools could help non-native English speakers refine scientific writing in English. Two of the earliest systematic reviews on this topic, e.g., Sallam M. [22] and Tustumi T. [23], have discussed the general use of ChatGPT in healthcare, both of which have concluded that further improvements and evaluations are required for ChatGPT to be a reliable tool for healthcare professionals, while highlighting its promising prospect in this field. Furthermore, it is interesting to notice that researchers working on the topic are actively interacting with each other. Gupta R. [28], which is published as a ‘letter to editor’, commented on the publication from Najafali D. [19], adding that the ethical concerns revolving around using ChatGPT in scientific writing still remain to be addressed, which is not sufficiently covered by the original publication. Similar to Gupta R. [28], Moreno E. [25] is also a comment on a previous publication i.e., Sallam M. [22], which, while acknowledging the contribution of Sallam M. [22] to raising the awareness among healthcare professionals of the inevitable prevalence of AI tools, such as ChatGPT in medical practice, pointed out the flaws of the work concerning the conformity to the PRISMA guideline. A response to the comment was published by the original authors [26], which addressed the critics and, at the same time, acknowledged the interest of Moreno E. et al. in their work. These comments and responses not only reflect a healthy interaction among researchers but also show that applying ChatGPT or other AI tools in healthcare is an active area of research.

Generally speaking, it is useful and necessary to discuss how ChatGPT will impact healthcare in abstract terms, as in Sallam M. [22] and Tustumi T. [23]. However, practical implementation of ChatGPT in healthcare practice requires detailed quantitative evaluations and careful consideration of the unique needs of different healthcare specialties [21]. This comprehensive approach ensures that ChatGPT can provide accurate, reliable, and useful assistance. To elaborate:

### Abstract Discussion of ChatGPT’s Impact on Healthcare

The works of Sallam M. [22] and Tustumi T. [23] have initiated discussions on how ChatGPT could influence healthcare at a conceptual level, which involves exploring how ChatGPT can assist in diagnosing diseases, providing medical advice, automating administrative tasks, refining and accelerating scientific writing. These discussions are crucial as they provide a theoretical framework for understanding the potential benefits and challenges of integrating AI into healthcare.

### Need for Detailed Quantitative Evaluations

While abstract discussions provide a theoretical understanding, they are often not enough for practical implementations. It’s necessary to conduct detailed quantitative evaluations of ChatGPT in various medical specialties and application scenarios. This could involve rigorous testing and validation of the AI model’s performance in real-world clinical settings. For instance, how accurately can ChatGPT diagnose a specific condition compared to a human doctor? How does its performance vary across different specialties like cardiology, neurology, or psychiatry, and how reliable is the performance? What legal and ethical issues and concerns are exposed, which should be addressed from a legislative and regulative perspective? Answering these questions is the prerequisite for the practical deployment of an AI tool, as discussed in [21].

### Consideration of Uniqueness and Special Requirements in Different Healthcare Specialties

Each medical specialty has its unique characteristics and requirements. For example, the type of language used, the complexity of cases, and the level of urgency can vary substantially. Therefore, for ChatGPT to be effectively implemented in healthcare practice, it must be tailored and optimized to meet these unique needs, which again requires individual evaluations as discussed above. This could be addressed by training the model on specialty-specific medical literature or fine-tuning it based on feedback from healthcare professionals in that specialty.

## Conclusion

Based on the systematic reviews about ChatGPT in the medical field in this contribution and the above discussions, we project that the number of systematic reviews will continue to increase, with more and more research on the evaluation of ChatGPT in a specific domain being published in the near future. However, it is important to note that, despite these reviews keeping track of the latest development of ChatGPT in healthcare, the quality and performance of the AI tool still depends on the underlying large language model, which needs to be continuously improved by researchers and practitioners in natural language processing (NLP). As healthcare professionals and researchers, our primary responsibility is to rigorously test the tool in our respective medical specialities in order to provide feedback on the actual capabilities of ChatGPT and expose its limitations and the ethical and legal concerns that arise. These in turn help NLP researchers to improve the language model and provide legislators the basis to formulate proper regulations. Summarized, we extracted the following three main challenges for ChatGPT in healthcare, but also medical NLPs in general, from our meta-review:

- **(1.) Dependence on the Underlying Language Model:** The quality and performance of ChatGPT are reliant on the underlying large language model, which is trained on vast amounts of text data and responsible for generating appropriate responses. Nevertheless, like any machine learning model, ChatGPT is prone to mistakes or failure given complex or ambiguous inputs. Therefore, continuous improvements by NLP researchers and practitioners are essential for fine-tuning the language model for a specific application. Furthermore, developing an artificial general intelligence (AGI) model with a wide spectrum of medical knowledge is a challenging but rewarding task.
- **(2.) Role of Healthcare Professionals and Researchers:** As users of ChatGPT in the medical field, healthcare professionals and researchers play an important role, who continuously provide feedback to NLP developers. Rigorous testing of the tool in their respective medical specialties is essential in providing valuable feedback on its actual capabilities in order to improve ChatGPT’s performance. The expert-in-the-loop process involves not only identifying applications that ChatGPT is good at, e.g., providing medical information or assisting with patient communication, but also helps to expose the limitations of ChatGPT, such as hallucination or inability to provide accurate information for complex medical questions. The feedback from healthcare professionals not only help NLP researchers to improve the language model but also provide legislators with a basis to formulate proper regulations.
- **(3.) Ethical and Legal Concerns:** Using AI tools like ChatGPT in healthcare can also inevitably raise ethical and legal issues, such as patient privacy and data security. A clear guideline on when and how ChatGPT should be used in a healthcare setting is required. Healthcare professionals, policymakers and regulators should work jointly to address these concerns.

To conclude, the development and use of AI tools like ChatGPT in healthcare is a collaborative effort that should involve NLP researchers, healthcare professionals and legislators. Each group has a crucial role to play in ensuring that these tools are (in this order) safe, beneficial and effective for patient care.

## Data Availability

All data produced in the present work are contained in the manuscript

## Acknowledgements

This work was supported by the REACT-EU project KITE (Plattform für KI-Translation Essen, https://kite.ikim.nrw/, EFRE-0801977) and FWF enFaced 2.0 (KLI 1044, https://enfaced2.ikim.nrw/). Behrus Puladi was funded by the Medical Faculty of the RWTH Aachen University in Germany as part of the Clinician Scientist Program. Furthermore, we acknowledge the Center for Virtual and Extended Reality in Medicine (ZvRM, https://zvrm.ume.de/) of the University Hospital in Essen, Germany.

## Disclaimer

For some parts of the paper, Microsoft’s Bing Chat was used to generate hints using customized prompts.

